# IL-6 and D-Dimer at Admission Predicts Cardiac Injury and Early Mortality during SARS-CoV-2 Infection

**DOI:** 10.1101/2021.03.22.21254077

**Authors:** Daoyuan Si, Beibei Du, Bo Yang, Lina Jin, Lujia Ni, Qian Zhang, Zhongfan Zhang, Mohammed Ali Azam, Patrick F.H. Lai, Stéphane Massé, Huan Sun, Xingtong Wang, Slava Epelman, Patrick R. Lawler, Ping Yang, Kumaraswamy Nanthakumar

**Affiliations:** Department of Cardiology, The Third Hospital of Jilin University, Jilin Provincial Molecular Biology Research Center for Precision Medicine of Major Cardiovascular Disease, Changchun, China; Institute of Organ Transplantation, Tongji Hospital, Tongji Medical College, Huazhong University of Science and Technology, Wuhan, China; Department of Epidemiology and Biostatistics, School of Public Health, Jilin University, Changchun, China; Department of Ultrasound, The Third Hospital of Jilin University, Changchun, China; The Hull Family Cardiac Fibrillation Management Laboratory, Toronto General Hospital, University Health Network, Toronto, Canada; Department of Hematology, The First Hospital of Jilin University, Changchun, China; Peter Munk Cardiac Centre, University Health Network, Toronto, Canada; The Ted Rogers Centre for Heart Research, Toronto, Canada; Interdepartmental Division of Critical Care Medicine, University of Toronto, Toronto, Canada

**Keywords:** COVID-19, Cardiac injury, immune activation, thrombotic activation, mechanism

## Abstract

**BACKGROUND:** We recently described mortality of cardiac injury in COVID-19 patients. Admission activation of immune, thrombotic biomarkers and their ability to predict cardiac injury and mortality patterns in COVID-19 is unknown.

**METHODS:** This retrospective cohort study included 170 COVID-19 patients with cardiac injury at admission to Tongji Hospital in Wuhan from January 29–March 8, 2020. Temporal evolution of inflammatory cytokines, coagulation markers, clinical, treatment and mortality were analyzed.

**RESULTS:** Of 170 patients, 60 (35.3%) died early (<21d) and 61 (35.9%) died after prolonged stay. Admission lab work that correlated with early death were elevate levels of interleukin 6 (IL-6) (p<0.0001), Tumor Necrosis Factor-a (TNF-a) (p=0.0025), and C-reactive protein (CRP) (p<0.0001). We observed the trajectory of biomarker changes after admission, and determined that early mortality had a rapidly increasing D-dimer, gradually decreasing platelet and lymphocyte counts. Multivariate and simple linear regression models showed that death risk was determined by immune and thrombotic pathway activation. Increasing cTnI levels were associated with those of increasing IL-6 (p=0.03) and D-dimer (p=0.0021). Exploratory analyses suggested that patients that received heparin has lower early mortality compared to those who did not (p =0.07), despite similar risk profile.

**CONCLUSIONS:** In COVID-19 patients with cardiac injury, admission IL-6 and D-dimer predicted subsequent elevation of cTnI and early death, highlighting the need for early inflammatory cytokine-based risk stratification in patients with cardiac injury.

**Condensed Abstract:** COVID-19 with cardiac injury is associated with worse survival. Admission activation of immune, thrombotic biomarkers and their ability to predict cardiac injury and mortality patterns in COVID-19 is unknown. This study proved that cardiac injury in these patients is closely related to the activation of immunological and thrombotic pathways and can be predicted by admission biomarkers of these pathways. This study supports the strategy of biomarker-guided, point-of-care therapy that warrants further studies in a randomized manner to develop anti-immune and anti-thrombotic treatment regimens in severe COVID-19 patients with cardiac injury.

## Introduction

Organ injury, including cardiac injury^1-5^, is common in patients with severe coronavirus disease 2019 (COVID-19) and is associated with worse survival^1, 6^. However, the underlying mechanism is yet elusive. The understanding of the mechanism that links COVID-19, which is caused by severe acute respiratory syndrome coronavirus-2 (SARS-CoV-2) infection with organ injury may reveal the upstream risk factors or biomarkers that may serve as potential therapeutic targets. Given the absence of a direct anti-viral treatment, there is an urgent need to develop such alternate therapeutic strategies. SARS-CoV-2 is thought to induce the activation of immune and thrombotic pathways, which may contribute to organ injury. To verify this hypothesis, we performed detailed longitudinal profiling of biomarkers of inflammation, coagulation, cardiac injury and mortality in 170 severe cases of COVID-19 at Tongji Hospital in Wuhan, China. Our analysis of these biomarkers indicated that immune and thrombotic activation in patients with COVID-19 determine the degree of post-admission cardiac injury and early death. Thus, the early detection and monitoring of these biomarkers of immune and thrombotic activation may help predict and prevent cardiac injury and death.

## Methods

### Patient population

Patient selection, data extraction and validation, definition of cardiac injury and ethical approval were detailed in our recent publication (CMAJ paper). Briefly we reported previously 1284 cases of COVID-19 pneumonia were diagnosed by computed tomography and transferred to Tongji Hospital between January 29 and March 8, 2020. Upon admission, the level of circulating cardiac Troponin I (cTnI) was measured in 1159 patients, of which 170 (14.7%) patients with cardiac injury, as indicated by elevated cTnI levels, were reviewed in this study. COVID-19 diagnosis was confirmed by laboratory testing performed according to the interim guidelines from the World Health Organization^7^. Blood specimens were collected within 72 h from admission, depending on each patient’s clinical situation, for testing as required by the treating physicians.

### Analysis of inflammatory and thrombotic cascade biomarkers

White blood cell (WBC), lymphocyte, and platelet counts (Sysmex XN9000, Japan), tumor necrosis factor-α (TNF-α), interleukin-6 (IL-6; Cobas e602, Germany), C-reactive protein (CRP; Cobas 8000, Germany), D-dimer, and fibrinogen levels, and international normalized ratio (INR), activated partial thromboplastin time (aPTT), and prothrombin time (Stago STA-R, France) were determined by Tongji Hospital Clinical Biochemistry Laboratory.

### Analysis the Effects of Anticoagulant Treatment

A portion of the study patients received heparin as anticoagulant venous thrombosis 4100 QD SC. To explore the potential preliminary effects of anticoagulant, we stratified the patients into two groups: the patients received or not received anticoagulant heparin. Mortality and need for invasive mechanical ventilation were compared in these two groups.

### Statistical analysis

Continuous variables were expressed as median (interquartile range [IQR]). Mann-Whitney tests were used for two-group comparisons, while Kruskal-Wallis test was used for comparisons between three groups. Categorical variables were expressed as proportions and percentages, and Fisher’s exact test was used to compare differences. Odds ratio (OR) and 95% confidence interval (CI) were separately calculated to predict in-hospital death using a multivariable regression model adjusted for age, sex, and comorbidities. A simple liner regression analysis was applied to examine the correlation between baseline biomarkers and peak cTnI level. Biomarker levels and dynamics over time were stratified by survival status, including (1) survival to discharge, (2) early (≤ 21 days) mortality, or (3) late (> 21 days) mortality. Statistical significance was defined as p ≤ 0.05. Data analysis was performed with GraphPad Prism 7.00 (San Diego, CA) software.

## Results

### Biomarkers characteristics of study patients at admission

Table 1 demonstrates the biomarker characteristics of study patients at admission; the patients were stratified into those discharged from hospital (survivors, n = 49) and those who died in hospital (non-survivors, n = 121). Demographic information of patients died and discharged were recorded in our recent reported [CMAJ 2020 In-press]. Levels of cTnI at admission were not different between survivors and non-survivors, while N-terminal pro b-type natriuretic peptide (NT-proBNP) levels were elevated in non-survivors at admission (median [IQR], 1043 [457.5, 4550] mg/L vs. 485 [219, 1106] mg/L for survivors, p < 0.001; **Table 1**). Coagulation abnormalities were observed in the non-survivor group upon admission, consistent with elevated D-dimer levels (median [IQR], 21 [11.16, 21] mg/L vs. 2.47 [0.92, 12.65] mg/L, p = 0.0005), increased PT, INR, and PTT, and reduced platelet count. Fibrinogen levels were not significantly different between the two groups. Inflammatory cytokine and CRP levels at admission were different between the survivors and non-survivors. Upon admission, CRP (median [IQR], 100.8 [60.6, 169.3] pg/mL vs. 51.1 [13.3, 77.6] pg/mL, p < 0.0001), TNF-α (median [IQR], 10.7 [7.8, 17.3] vs. 8 [5, 11.8] pg/mL, p = 0.02), and IL-6 (median [IQR], 68.01 [28.69, 192.6] vs. 17.27 [7.08, 41.1] pg/mL, p < 0.0001) levels were higher in non-survivors than in survivors **(Table 1)**. According to these results, immune dysregulation was evident at admission.

**Table 1.**
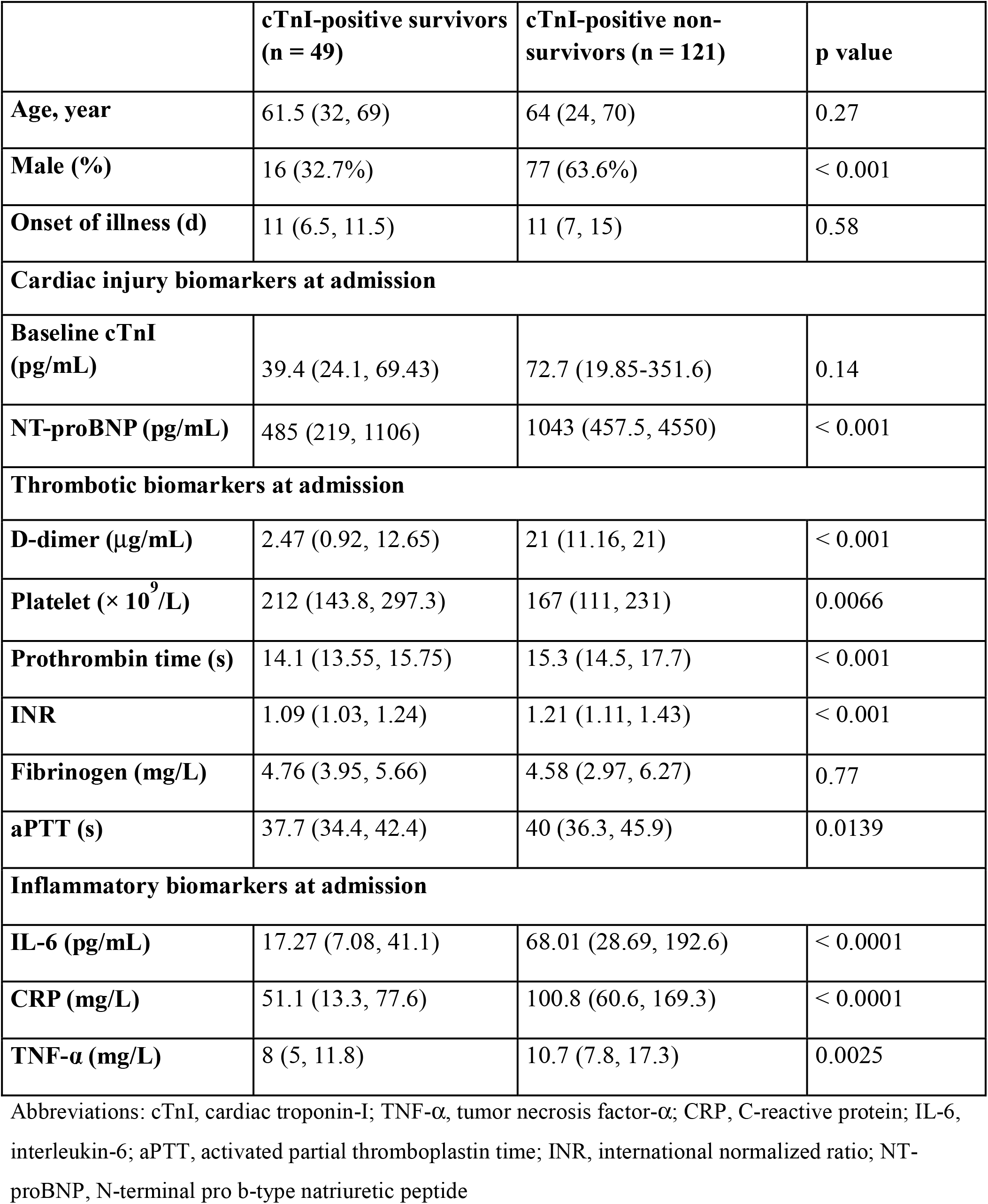
Demographics, Cardiac Injury, and Biomarker Characteristics of COVID-19 Patients.

### Analysis of cardiac injury biomarkers

We tracked cTnI levels over 30 days and found them to be markedly increased in patients who eventually died **(Figure 1A)**. A drop in median cTnI levels in patients who died during hospitalization suggested a survival bias between patients who died before day 21 versus those that died later. We stratified our analysis by comparing those who died between day 6 of symptom onset, the earliest time correlating near to admission, and day 21 (early mortality cohort) with patients who died after day 21 (late mortality cohort). In the early mortality cohort, cTnI levels were high, and patients who died late had lower cTnI levels similar to those of survivors **(Figure 1B)**. A similar trend was observed in NT-proBNP levels, which were higher in patients from the early mortality group than in survivors and late mortality groups **(Figure 1C)**. These data suggest that an increasing trajectory of cTnI and NT-proBNP (beyond admission levels) predicted early mortality and that patients who died after 21 days from illness onset had cTnI and NT-proBNP levels similar to those of survivors.

**Figure 1.** Cardiac injury biomarkers in patients with COVID-19. **A**. Temporal changes in cTnI levels among survivors and dead patients affected with COVID-19. **B**. Temporal changes in cTnI levels in discharged patients and those with early and late mortality. **C**. Temporal changes in NT-proBNP levels in discharged patients and those with early and late mortality. *p □ 0.05

### Evaluation of immune biomarkers

We focused on the early phases of the disease and observed that CRP levels markedly improved over hospitalization in survivors but remained elevated in those with early mortality **(Figure 2A)**. Elevated WBC and decreased lymphocyte counts correlated with early mortality **(Figure 2B-C)**. While TNF-α levels were elevated only at admission, IL-6 levels remained high in those with early mortality **(Figure 2D-E)**. In comparison with the late mortality group, the patients from the early mortality group had higher levels of IL-6 (median [IQR]: 23.86 [10.76, 85.78] vs. 179.4 [37.25, 736] vs. 15.23 [2.84, 41.6] pg/mL in discharge vs. early death vs. late death groups, respectively; p = 0.02), TNF-α (8 [7.3, 11.8] vs. 19 [12.45, 28.58] vs. 7.75 [4.78, 9.98] pg/mL in discharge vs. early death vs. late death groups, respectively; p = 0.003), and CRP (65.35 [23.6, 94.35] vs. 138.8 [111, 191] vs. 39.25 [36.61, 58.8] mg/L in discharge vs. early death vs. late death groups, respectively; p = 0.0004) **(Figure 2F)**. These data suggest that early death in patients with COVID-19 was associated with the activation of the inflammatory immune cascade, a phenomenon already evident at admission.

**Figure 2.** Biomarkers of immune activation in patients with COVID-19. In comparison with survivors, patients with early mortality showed marked increase in CRP levels **(A)** over hospitalization. Elevated WBC and decreased lymphocyte counts correlated with early mortality (**B, C)**. TNF-α levels were elevated only at admission **(D)**. IL-6 levels remained elevated in those with early mortality (**E)**. IL-6, TNF-α, and CRP levels at admission were compared between survivors and patients with early mortality (within 21 days) and late mortality (after day 21). IL-6, TNF-α, and CRP levels were elevated only in patients who died early (**F**). *p □ 0.05

### Evaluation of thrombotic biomarkers

The most notable abnormalities among thrombotic markers were the rapid increase in D-dimer levels from day 9 after illness onset along with a rapid decrease in platelet counts beginning on day 12 among non-survivors **(Figure 3A-B)**. aPTT, prothrombin time, and INR increased over time in the early mortality cohort **(Figure 3C-E)**, whereas no differences in fibrinogen levels were observed over the time course between non-survivors and survivors **(Figure 3F)**.

**Figure 3.** Activation profile of thrombosis biomarkers in patients with cardiac injury. Prominent increase in D-dimer levels occurred since day 9 from illness onset and platelet counts rapidly decreased at day 12 from illness onset in patients who died early as compared to those in survivors (**A, B**). INR, prothrombin time, and aPTT increased over time in the early mortality cohort (**C-E**). No differences in fibrinogen levels were observed over the time course in two groups (**F**). *p □ 0.05

### Biomarkers predictive of cardiac injury and death

**Table 2A** describes the linear regression analysis results for the biomarkers of cardiac injury based on peak cTnI values after adjusting for age, sex, history of coronary heart disease, hypertension and diabetes mellitus. Baseline IL-6 (β 0.243 [0.018, 0.468], p = 0.03) and D-dimer (β 0.27 [0.042, 0.50], p = 0.0021) predicted cardiac injury, suggestive of the ability of early immune and thrombotic activation to predict cardiac injury. We investigated the biomarkers that could predict mortality in a multi-variate model after adjusting the aforementioned demographics/comorbidities and found that the level of D-dimer (OR 4.202 [2.15, 8.20], p < 0.001), IL-6 (OR 8.382 [3.28, 21.40], p < 0.001), and TNF-α (OR 9.012 [1.53, 52.94], p = 0.015), but not cTnI levels at admission predicted mortality **(Table 2B)**.

**Table 2.** Multivariable Regression Analysis of Death and Peak cTnI.

| **A. Simple linear regression prediction of peak cTnI with D-dimer, IL-6, and TNF-α** |  |
| --- | --- |
|  | **$\beta^*$ (95% CI\*)** |
| **Log D-dimer** | 0.271 (0.042, 0.500) |
| **Log IL-6** | 0.243 (0.018, 0.468) |
| **Log TNF-α** | 0.125 (−0.498, 0.749) |
| **B. Multivariate logistic regression analysis of death from cardiac injury, thrombosis, and immunological parameters at admission** |  |
|  | **OR\* (95% CI\*)** |
| **Log baseline cTnI** | 1.35 (0.83, 2.20) |
| **Log D-dimer** | 4.202 (2.15, 8.20) |
| **Log IL-6** | 8.382 (3.28, 21.40) |
| **Log TNF-α** | 9.012 (1.53, 52.94) |

\*Adjusted by age, sex, coronary heart disease, and hypertension
Abbreviations: TNF-α , tumor necrosis factor-α ; IL-6, interleukin-6; cTnI, cardiac troponin-I; CI, confidence interval

**Table 3.** Demographics on exploratory heparin treatment.

|  | **Heparin on in discharged and early death(n=18)** | **Heparin off in discharged and early death(n=91)** | **P value** |
| --- | --- | --- | --- |
| **Age, yrs** | 70.5 (65.5, 79) | 70 (64, 78) | 0.80 |
| **Male (%)** | 7 (38.9) | 45 (49.5) | 0.45 |
| **Onset of illness (d)** | 7.5 (4.75, 14.25) | 10 (6, 14) | 0.40 |
| **Past medical history** |  |  |  |
| **Pre-existing CHD (%)** | 2 (11.1%) | 21 (23.1%) | 0.35 |
| **Hypertension (%)** | 7 (38.9) | 45 (49.5) | 0.45 |
| **Diabetes (%)** | 5 (27.8%) | 24 (26.4%) | >0.99 |
| **Admission Cardiac Injury Biomarkers** |  |  |  |
| **Baseline cTnI (pg/ml)** | 30.6 (19.1, 121.2) | 46.6 (24.5, 118) | 0.35 |
| **NT-proBNP (pg/ml)** | 723 (320.3, 2075) | 808 (406.5, 4800) | 0.42 |
| **Admission Thrombotic Biomarkers** |  |  |  |
| **D-dimer ( ug/ml )** | 8.16 (1.12, 15.1) | 2.11 (1.06, 16.6) | 0.54 |
| **Admission Inflammatory Biomarkers** |  |  |  |
| **IL-6 (pg/ml)** | 44.6 (29.6, 85.8) | 39.6 (9.8, 117.4) | 0.62 |
| **CRP (mg/L)** | 72.3(44.9, 135.3) | 81.6 (40.2, 135.4) | 0.78 |
| **TNFα(mg/L)** | 11(8.6, 18.2) | 9 (5.9, 14.8) | 0.35 |

### Effects of Incidental Heparin Therapy on Cardiac Injury

VTE prophylaxis heparin treatment was initiated during hospitalization in 18 patients in discharged and early death group. In-hospital adverse outcomes (mortality or mechanical ventilation support) were compared in the 18 heparin-treated patients and 91 patients who were not treated with heparin. Baseline characteristics were compared between heparin on and heparin-off groups, which showed no significant differences. Mortality has a trend of decrease in heparin-treated patients compared with those not treated with heparin (33.3% vs 59.3%, p=0.07). There was no difference in the rates of mechanical ventilation (35.7% vs 32.6%, p= 0.99). **(Figure 4)**

**Figure 4.** Exploratory effect of heparin treatment on mortality and IMV in cardiac injury. Exploratory heparin treatment has a trend of decrease in mortality (p=0.07) and IMV need (p=0.99). IMV: invasive mechanical ventilation.

## Discussion

In the present study, levels of cTnI continued to increase during hospitalization, and peak cTnI values were several folds higher prior to death. The levels of the thrombotic biomarker D-dimer, inflammatory cytokines IL-6 and TNF-α, and inflammatory marker CRP significantly increased early during hospitalization in COVID-19 patients who died as compared with survivors. Peak cTnI levels correlated with the prior increase in IL-6 and TNF-α levels, suggesting that activation of immune and thrombotic pathways was preceded by cardiac injury. Our study provides a clear understanding of the pathogenic interaction between immune activation, coagulation, and cardiac injury in COVID-19 infection and may help identify novel therapeutic targets to mitigate poor outcomes in severe COVID-19 cases. The interactions we have demonstrated may provide opportunities to improve clinical care and design therapy protocols for patients with severe COVID-19.

### Immune dysregulation and coagulation disturbance in patients with COVID-19

Emerging evidence of lung injury from earlier pilot studies is associated with the increased levels of IL-6 in COVID-19 patients with deteriorated lung functions and increased need for mechanical ventilation^8-10^. Several cytokines and their receptors such TNF-α, IL-2, IL-2R, and IL-8 were elevated (single time point) in patients with more severe symptoms and positively correlated with the viral load and increased cTnI levels^11^. Our results demonstrate that IL-6 and TNF-α are the key biomarkers reflective of the cardiac status of patients with COVID-19 early during hospitalization; these markers may highlight patients at risk before the rise in cTnI level. While immune activation is suspected to be another mechanism accompanied with respiratory complications in COVID-19, its exact association with cardiac injury is unclear. Emerging evidence suggests the dysregulation, not simply overactivation, of immune response. For instance, a detailed immuno-phenotyping study of 56 patients with COVID-19 showed that those with severe respiratory failure exhibited a marked decrease in the level of human leukocyte antigen (HLA-DR), which appears to be driven by sustained IL-6 and TNF-α production^12^. Interestingly, this pattern is not observed in severe bacterial pneumonia. Moreover, while inflammatory cytokines dominate, the relative absence of a type I interferon response needed for viral clearance^8, 13, 14^ suggests a pattern specific for SARS-CoV-2.

Whether alone or in combination, various inflammatory factors are capable of inducing cardiac injury. The SARS-CoV-2–specific inflammatory milieu may lead to cardiac dysfunction. Viral myocarditis or the recently described viral infection of endothelial cells^15^ potentially leading to microvascular injury are other plausible mechanisms. While IL-6 levels were elevated in patients with a risk of early death, extremely high IL-6 levels noted in our study may be directly related to uncontrolled viremia observed elsewhere^16^. Receptor antagonists of these cytokines may attenuate the inflammatory action and prevent the escalation of the cytokine storm, thereby suppressing the overwhelming response of SARS-CoV-2 on the heart before development of uncontrolled viremia. Thus, immune and thrombotic cascade modulation may provide a direct means and a potentially life-saving strategy during aggressive antiviral treatments in high-risk patients and prevent disease progression toward mortality.

### Evidence of hypercoagulability in patients with COVID-19

Coagulation derangements in patients with COVID-19 are strongly associated with poor clinical outcomes, and various lines of evidence suggest that the prothrombotic state may be causal. Elevated D-dimer may be a biomarker of this pathway^1^, although D-dimer elevation in COVID-19 may be multi-factorial^17^. In a series of 183 patients with COVID-19, the non-survivors (11%) exhibited elevated D-dimer and fibrin degradation products; 15 non-survivors met the criteria for disseminated intravascular coagulation (DIC), whereas only 1 survivor developed DIC^18^. Similar derangements were documented in a separate case series of 94 patients with COVID-19 ^19^. Markers of DIC correlate with clinical deterioration, including ischemic injury of fingers and toes^20^. Macrovascular embolic events may occur with underappreciated frequency^21^. Acute pulmonary embolism and deep venous thrombosis had been reported in patients with SARS-CoV-2 infection^22, 23^. The limited autopsy data suggest a constellation of pathological findings, including thrombus in pulmonary microvessels^24^. Therefore, D-dimer levels were higher and tended to rise to a greater extent among non-survivors. Thus, patients with COVID-19 may exhibit an underlying pro-thrombotic state.

### Cardiac injuries and thrombotic pathways

Previous studies have reported the progressive rise in D-dimer and cTnI levels in patients who subsequently died versus survivors^1^. Patients with D-dimer values higher than 1 mcg/mL at admission were at increased risk of in-hospital death (adjusted OR: 18.42; 95% CI: 2.64, 128.55; p = 0.0033). The comparison between 113 COVID-19 non-survivors and 161 survivors by Chen and colleagues showed that cTnI and D-dimer levels were markedly higher in non-survivors (median cTnI level: 40.8 [IQR: 14.7, 157.8] vs. 3.3 [IQR: 1.9, 7.0] pg/mL; median D-dimer level: 4.6 [IQR: 1.3, 21.0] vs. 0.6 [IQR: 0.3, 1.3] pg/mL)^6^. These biomarkers may be a valuable aid for risk stratification and guidance on resource allocation among hospitalized patients and to design an active cardiac surveillance strategy for COVID-19 management^25^.

### Limitations

To prevent the spread of COVID-19, the assessment of ventricular functions by echocardiography to determine cardiomyopathy was not routinely performed. Therefore, we were unable to correlate the rising levels of cTnI to cardiac contractile function. We only reviewed the data on cTnI-positive patients. The collection of biomarkers testing was not formatted and left to the discretion of the COVID team; this may introduce a bias. For instance, of the 170 patients with elevated cTnI levels, 38 had no data on baseline IL-6 or D-dimer levels owing to the early experience of pandemic medical response.

### Conclusion

Cardiac injury in patients with COVID-19 is closely related to the activation of immunological and thrombotic pathways and can be predicted by admission biomarkers of these pathways. This study supports the strategy of biomarker-guided, point-of-care therapy that warrants further studies in a randomized manner to develop anti-immune and anti-thrombotic treatment regimens in severe COVID-19 patients with cardiac injury.

## Data Availability

The datasets used and/or analysed during the current study are available from the corresponding author on reasonable request.

## Acknowledgements

This research was supported by grants from the Excellent Youth Foundation of Science and Technology of Jilin Province (No.20180520054JH) and the “13^th^ Five-Year” Science Project of the Department of Education of Jilin Province (No. JJKH20190062KJ).

## References

1. Zhou F, Yu T, Du R, Fan G, Liu Y, Liu Z, Xiang J, Wang Y, Song B, Gu X, Guan L, Wei Y, Li H, Wu X, Xu J, Tu S, Zhang Y, Chen H and Cao B. Clinical course and risk factors for mortality of adult inpatients with COVID-19 in Wuhan, China: a retrospective cohort study. The Lancet. 2020.

2. Shi S, Qin M, Shen B, Cai Y, Liu T, Yang F, Gong W, Liu X, Liang J, Zhao Q, Huang H, Yang B and Huang C. Association of Cardiac Injury With Mortality in Hospitalized Patients With COVID-19 in Wuhan, China. JAMA Cardiology. 2020.

3. Ruan Q, Yang K, Wang W, Jiang L and Song J. Clinical predictors of mortality due to COVID-19 based on an analysis of data of 150 patients from Wuhan, China. Intensive Care Med. 2020.

4. Wang D, Hu B, Hu C, Zhu F, Liu X, Zhang J, Wang B, Xiang H, Cheng Z, Xiong Y, Zhao Y, Li Y, Wang X and Peng Z. Clinical Characteristics of 138 Hospitalized Patients With 2019 Novel Coronavirus–Infected Pneumonia in Wuhan, China. JAMA. 2020;323:1061–1069.

5. Chen N, Zhou M, Dong X, Qu J, Gong F, Han Y, Qiu Y, Wang J, Liu Y, Wei Y, Xia J, Yu T, Zhang X and Zhang L. Epidemiological and clinical characteristics of 99 cases of 2019 novel coronavirus pneumonia in Wuhan, China: a descriptive study. Lancet (London, England). 2020;395:507–513.

6. Chen T, Wu D, Chen H, Yan W, Yang D, Chen G, Ma K, Xu D, Yu H, Wang H, Wang T, Guo W, Chen J, Ding C, Zhang X, Huang J, Han M, Li S, Luo X, Zhao J and Ning Q. Clinical characteristics of 113 deceased patients with coronavirus disease 2019: retrospective study. BMJ (Clinical research ed). 2020;368:m1091.

7. WHO. Clinical management of severe acute respiratory infection when Novel coronavirus (nCoV) infection is suspected: interim guidance. https://www.who.int/publications-detail/clinical-management-of-severe-acute-respiratory-infection-whennovel-coronavirus-(ncov)-infection-is-suspected. 2020.

8. Lagunas-Rangel FA and Chavez-Valencia V. High IL-6/IFN-gamma ratio could be associated with severe disease in COVID-19 patients. J Med Virol. 2020.

9. Buonaguro FM, Puzanov I and Ascierto PA. Anti-IL6R role in treatment of COVID-19-related ARDS. J Transl Med. 2020;18:165.

10. Conti P, Ronconi G, Caraffa A, Gallenga CE, Ross R, Frydas I and Kritas SK. Induction of proinflammatory cytokines (IL-1 and IL-6) and lung inflammation by Coronavirus-19 (COVI-19 or SARS-CoV-2): anti-inflammatory strategies. J Biol Regul Homeost Agents. 2020;34.

11. Liu Y, Liao W, Wan L, Xiang T and Zhang W. Correlation Between Relative Nasopharyngeal Virus RNA Load and Lymphocyte Count Disease Severity in Patients with COVID-19. Viral Immunol. 2020.

12. Giamarellos-Bourboulis EJ, Netea MG, Rovina N, Akinosoglou K, Antoniadou A, Antoonakos N and Koutsoukou A. COMPLEX IMMUNE DYSREGULATION IN COVID-19 PATIENTS WITH SEVERE RESPIRATORY FAILURE. Cell. 2020.

13. O’Brien TR, Thomas DL, Jackson SS, Prokunina-Olsson L, Donnelly RP and Hartmann R. Weak Induction of Interferon Expression by SARS-CoV-2 Supports Clinical Trials of Interferon Lambda to Treat Early COVID-19. Clin Infect Dis. 2020.

14. Chu H, Chan JF, Wang Y, Yuen TT, Chai Y, Hou Y, Shuai H, Yang D, Hu B, Huang X, Zhang X, Cai JP, Zhou J, Yuan S, Kok KH, To KK, Chan IH, Zhang AJ, Sit KY, Au WK and Yuen KY. Comparative replication and immune activation profiles of SARS-CoV-2 and SARS-CoV in human lungs: an ex vivo study with implications for the pathogenesis of COVID-19. Clin Infect Dis. 2020.

15. varga Z, Flammer A, Steiger P, M H, R A, As Z, Mr M R S F R and H M. Endothelial cell infection and endotheliitis in COVID-19. Lancet. 2020.

16. Wu CI, Postema PG, Arbelo E, Behr ER, Bezzina CR, Napolitano C, Robyns T, Probst V, Schulze-Bahr E, Remme CA and Wilde AAM. SARS-CoV-2, COVID-19 and inherited arrhythmia syndromes. Heart Rhythm. 2020.

17. van de Veerdonk F, Netea MG, van Deuren M, van der Meer Jwm, de Mast Q, Bruggemann RJ and van der Hoeven H. Kinins and cytokines in COVID-19: a comprehensive pathophysiological approach. Preprints. 2020:2020040023.

18. Tang N, Li D, Wang X and Sun Z. Abnormal coagulation parameters are associated with poor prognosis in patients with novel coronavirus pneumonia. J Thromb Haemost. 2020;18:844–847.

19. Lippi G and Plebani M. Laboratory abnormalities in patients with COVID-2019 infection. Clinical chemistry and laboratory medicine. 2020.

20. Lin L, Lu L, Cao W and Li T. Hypothesis for potential pathogenesis of SARS-CoV-2 infection-a review of immune changes in patients with viral pneumonia. Emerging microbes & infections. 2020;9:727–732.

21. Poston JT, Patel BK and Davis AM. Management of Critically Ill Adults With COVID-19. JAMA. 2020.

22. Danzi GB, Loffi M, Galeazzi G and Gherbesi E. Acute pulmonary embolism and COVID-19 pneumonia: a random association? Eur Heart J. 2020.

23. Wichmann D, Sperhake JP, Lutgehetmann M, Steurer S, Edler C, Heinemann A, Heinrich F, Mushumba H, Kniep I, Schroder AS, Burdelski C, de Heer G, Nierhaus A, Frings D, Pfefferle S, Becker H, Bredereke-Wiedling H, de Weerth A, Paschen HR, Sheikhzadeh-Eggers S, Stang A, Schmiedel S, Bokemeyer C, Addo MM, Aepfelbacher M, Puschel K and Kluge S. Autopsy Findings and Venous Thromboembolism in Patients With COVID-19: A Prospective Cohort Study. Annals of internal medicine. 2020.

24. Yao XH, Li TY, He ZC, Ping YF, Liu HW, Yu SC, Mou HM, Wang LH, Zhang HR, Fu WJ, Luo T, Liu F, Chen C, Xiao HL, Guo HT, Lin S, Xiang DF, Shi Y, Li QR, Huang X, Cui Y, Li XZ, Tang W, Pan PF, Huang XQ, Ding YQ and Bian XW. A pathological report of three COVID-19 cases by minimally invasive autopsies. Zhonghua Bing Li Xue Za Zhi. 2020;49:E009.

25. Chapman AR, Bularga A and Mills NL. High-Sensitivity Cardiac Troponin Can Be An Ally in the Fight Against COVID-19. Circulation. 2020.

